# A Natural Language Processing Tool to Extract Quantitative Smoking Status from Clinical Narratives

**DOI:** 10.1101/2020.10.30.20223511

**Authors:** Xi Yang, Hanyuan Yang, Tianchen Lyu, Shuang Yang, Yi Guo, Jiang Bian, Hua Xu, Yonghui Wu

## Abstract

This study presents a natural language processing (NLP) tool to extract quantitative smoking information (e.g., Pack-Year, Quit Year, Smoking Year, and Pack per Day) from clinical notes and standardized them into Pack-Year unit. We annotated a corpus of 200 clinical notes from patients who had low-dose CT imaging procedures for lung cancer screening and developed an NLP system using a two-layer rule-engine structure. We divided the 200 notes into a training set and a test set and developed the NLP system only using the training set. The experimental results on the test set showed that our NLP system achieved the best F1 scores of 0.963 and 0.946 for lenient and strict evaluation, respectively.

**Note:** Accepted as a presentation at the 2020 IEEE International Conference on Healthcare Informatics (ICHI) Workshop on Health Natural Language Processing (HealthNLP 2020).

https://ohnlp.github.io/HealthNLP2020/healthnlp2020#.

## I. Introduction

Tobacco use is a significant risk factor associated with numerous diseases such as cancers and cardiovascular diseases [1]. Electronic health record (EHR) systems capture smoking status of patients in both structured EHR data and narrative clinical notes. Yet, detailed smoking information is more likely embedded in clinical narratives. Therefore, natural language processing (NLP) systems have been developed to extract categorical smoking status of patients (e.g., non-smoker, past smoking, current smoker) from clinical notes, such as the smoking status detection modules in the clinical Text Analysis and Knowledge Extraction System (cTAKES) [2] and the Clinical Language Annotation, Modeling, and Processing Toolkit (CLAMP) [3]. However, in clinical practices, detailed quantitative tobacco use information is often required. For example, the United States Preventive Services Task Force (USPSTF) recommends annual screening for lung cancer with low-dose computed tomography (CT) in high-risk adults: (1) aged 55 to 80 years, (2) who have a 30-pack-year smoking history, and (3) currently smoke or have quit smoking within the past 15 years. Being able to extract the quantitative (i.e., pack-year) smoking information is critical to assess one’s lung cancer risk.

This study developed a clinical NLP tool to extract quantitative smoking information from clinical notes to support studies that require quantitative smoking information. We constructed a clinical corpus with smoking information manually annotated using clinical notes from the University of Florida (UF) Health Integrated Data Repository (IDR)—a clinical data warehouse, and developed an NLP tool to systematically extract the quantitative smoking information (e.g., pack-day, pack-year) from clinical narratives. We evaluated the NLP tool using standard NLP evaluation metrics and further compared the NLP extracted smoking information with the structured EHR data to assess differences.

## II. Material and Methods

### A. Data sets

In this study, we extracted a corpus from the UF Health IDR based on a cohort of 3,080 patients who received lung cancer screening between 2012 and 2019. We filtered the corpus with a set of keywords derived from “*smoke*” and “*tobacco*” to remove smoking-unrelated clinical notes. With the rest of the notes, we randomly selected 200 notes for annotation.

Two annotators (TL and SY) labeled the mentions of quantitative smoking information, including Pack-Year (a measurement unit for how much a person has smoked during a period of time, 1 pack-year means a person smoked 1 pack a day for one year [4]), Quit Year, Pack per Day, and Smoking Year. Based on 50 overlapped notes, we assessed the inter-annotator agreement using the Cohen’s kappa measurement [5]. We only focused on the quantitative smoking information pertaining to a specific patient and excluded general or instructional information such as the smoking information mentioned in guidelines (e.g. “*we discussed guidelines including patients with greater or equal than 30 pack-year smoking history*”). The 200 notes were divided into a training set of 160 notes and a test set of 40 notes. The study was approved by the University of Florida Institutional Review Board.

### B. Development of the NLP tool

We approached this information extraction task using a rule-engine approach. To better capture different writing patterns of the quantitative smoking information, we developed a two-layer rule-engine. The first layer consists of lexicons defined using regular expressions, which were later used to define high-level rules in the second layer. The developer (HY) designed lexicons and optimized rules by observing the smoking information annotated in the training set. The test set was not used during the system development and optimization. The system with the optimized performance on the training set was used to predict on the test set for final evaluation.

### C. Experiment and Evaluation

We evaluated the smoking information extracted by our NLP system using the manual annotation as a gold standard. The micro-averaged precision, recall and F1 score were used for evaluation. Similar to the named entity recognition (NER) task, we calculated both the strict and lenient scores using the official evaluation script released by 2019 n2c2 challenge organizers.

## III. Results

The inter-annotator agreement score between the two annotations was 0.91 (Cohen’s kappa). Our NLP system achieved the best lenient and strict F1 score of 0.963 and 0.946, respectively. Table I shows the evaluation scores for each category on the test set. We also compared the NLP extracted smoking information with the information documented in structured EHR for 10 patients to assess the consistency on the patient/year level. For each patient, we collected the yearly Pack-Year information from the structured EHR and compared them with the corresponding information extracted from clinical notes. A total number of 81 data points (on the patient/year level) were observed from the 10 patients. The comparison results showed that NLP is consistent with the structured EHR for 44 patient/year data points (54%), inconsistent for 5 patient/year data points (6%). For 10 patient/year data points (12%), we can only get the Pack-Year information from notes, whereas, for another 10 patient/year data points (12%) we only can get information from structured EHR. There 12 patient/year data points (15%) missing from both NLP and structured EHR.

**Table I.**
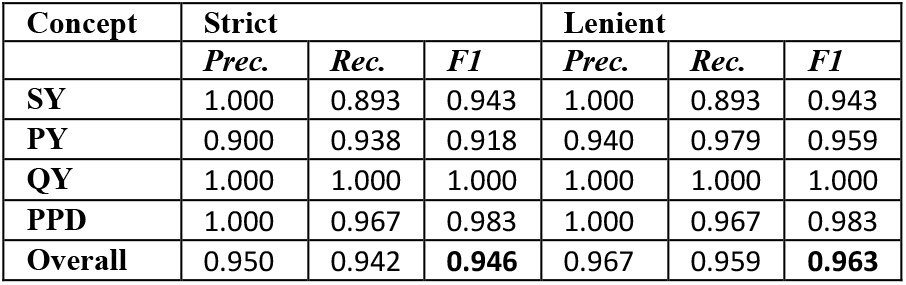
Evaluation results on test set.

## IV. Discussion and Conclusion

This study presents a simple clinical NLP tool to extract quantitative smoking information from clinical narratives. We compared the smoking information from notes with the structured EHR data and demonstrated that the quantitative smoking information from clinical notes is a good supplement to the information documented in structured EHRs.

## Data Availability

Not a public dataset

## ACKNOWLEDGMENT

This study was supported by a grant from the National Cancer Institute, 1R01CA246418, a Patient-Centered Outcomes Research Institute® (PCORI®) Award (ME-2018C3-14754), and the Cancer Informatics and eHealth core jointly supported by the UF Health Cancer Center and the UF Clinical and Translational Science Institute. The content is solely the authors’ responsibility and does not necessarily represent the official views of the funding institutions.

